# Mudan Granules for Diabetic Peripheral Neuropathy: A Systematic Review of Preclinical and Clinical Evidence

**DOI:** 10.1101/2023.05.30.23290766

**Authors:** Jianlong Zhou, Lv Zhu, Yadi Li

**Affiliations:** Department of Traditional Chinese Medicine, People’s Hospital of Deyang City, Deyang, China; Department of Integrative Medicine, West China Hospital, Sichuan University, Chengdu, China

## Abstract

**Background:** Mudan granules has been used in China to treat diabetic peripheral neuropathy (DPN), but there is a lack of systematic review of reports in this area. The aim of this systematic review was to evaluate the efficacy and safety of Mudan granules in the treatment of diabetic peripheral neuropathy.

**Methods:** Initial studies were searched from PubMed, Embase, Cochrane, China National Knowledge Infrastructure (CNKI), VIP database, Wanfang electronic databases. The Cochrane Risk of Bias tool was used to evaluate the risk of bias. The meta-analysis was performed by Stata 16.0 software. For dichotomous and continuous outcomes, the relative risk (RR) and standardized mean difference (SMD) with 95% confidence interval (CI) were conducted, respectively.

**Results:** 51 randomized controlled trials (RCTs) involving 5,416 patients were included. In the meta-analysis, compared with routine treatment (RT) alone, Mudan granules plus RT reduced Toronto clinical scoring system (TCSS) score (SMD, −0.52; 95% CI, −0.66 to −0.38; *P* < 0.01), total symptoms score (TSS) (SMD, −1.44; 95% CI, −2.88 to −0.00; *P* = 0.05), serum homocysteine (Hcy) levels (SMD, - 3.84; 95% CI, −5.99 to −1.70; *P* < 0.01), serum high sensitive C-reactive protein (hs-CRP) levels (SMD, −1.68; 95% CI, −3.29 to −0.08; *P* = 0.04), and improved total clinical efficacy (RR, 1.23; 95% CI, 1.19 to 1.27; *P* < 0.01) and serum superoxide dismutase (SOD) levels (SMD, 1.54; 95% CI, 1.13 to 1.95; *P* < 0.01). Besides, Mudan granules presented an adjuvant efficacy on median motor nerve conduction velocity (SMD, 1.61; 95% CI, 1.16 to 2.07), median sensory nerve conduction velocity (SMD, 1.73; 95% CI, 1.26 to 2.20), common peroneal motor nerve conduction velocity (SMD, 1.48; 95% CI, 1.10 to 1.86), common peroneal sensory nerve conduction velocity (SMD, 1.57; 95% CI, 1.23 to 1.92), tibial motor nerve conduction velocity (SMD, 1.34; 95% CI, 0.82 to 1.87), and tibial sensory nerve conduction velocity (SMD, 1.03; 95% CI, 0.86 to 1.20). In terms of adverse events, there was no statistically significant difference between the trial group and the control group (*P* =0.87). Five preclinical studies were also retrieved for the study. Animal studies have shown that Mudan granules have anti-oxidative stress effects and could reduce the inflammatory response. It may improve peripheral nerve injury in diabetic rats by modulating the TLR4/MyD88/NF-κB pathway, TLR4/p38 MAPK pathway and PI3K/AKT pathway.

**Conclusions:** Mudan granules presented an adjuvant efficacy on patients with DPN and could improve the oxidative stress and inflammatory levels in preclinical models. However, high-quality original studies are needed to further prove the evidence.

## 1. Introduction

Diabetic peripheral neuropathy is one of the most common chronic complications of diabetes, occurring in approximately 50% of people with diabetes(1). It is also an important cause of lower limb amputation and disabling neuropathic pain. It causes great suffering to patients in their daily lives and places a heavy economic burden on the healthcare system and society as a whole(2). Previous studies have reported prevalence rates of DPN ranging from 6% to 51%, with this variation being related to the population studied(3). 50% of people with DPN are usually asymptomatic in the early stages(4). There are also no simple markers for the early detection of DPN in routine clinical practice. DPN is defined internationally as “diabetic patients accompanied by signs and/or symptoms associated with peripheral nerve dysfunction after other causes have been excluded”(4). This definition suggests that the diagnosis of DPN is comprehensive and exclusive. A diagnosis of DPN requires 3 elements to be met. Firstly, it is necessary to have definite diabetes. Secondly, there must be clinical evidence of the presence of peripheral neuropathy (signs or symptoms), and/or evidence of electrophysiological examination. Finally, other causes of peripheral neuropathy need to be ruled out by relevant laboratory tests. The Toronto Consensus recommends the use of abnormal nerve conduction studies (NCS) with signs or symptoms to diagnose DPN. The NCS is considered to be the most important basis for the diagnosis of large fiber neuropathy. Screening for DPN involves a foot examination and history taking for neurological symptoms, as well as screening tests such as the 128 Hz tuning fork vibration sensing test and the 10 g (Semmes-Weinstein) monofilament test(2). However, these tests are unable to detect lesions in small nerve fibers. Skin biopsy is considered the reference for identifying small fiber neuropathy. Nevertheless, mass screening and repeat biopsies are not feasible(5). Therefore, the diagnosis of DPN, the determination of global prevalence and incidence remains a challenging task.

Patients with DPN usually present with symmetrical pain in the extremities, especially in the distal extremities. The most typical manifestation is abnormal sensation in the “glove-and-stocking” distribution(6). Current clinical management focuses on glycemic control, foot care and pain management(3). Strict glycemic control may stop the progression of diabetic neuropathy, but there is no evidence that glycemic control relieves pain in DPN. For the management of pain in DPN, major international clinical guidelines recommend several symptomatic treatment protocols(7). First-line treatment options include tricyclic antidepressants, serotonin-norepinephrine reuptake inhibitors and anticonvulsants that act on calcium channels. Pregabalin, gabapentin and duloxetine are the usual first-line drugs. However, tricyclic antidepressants could cause dry mouth, orthostatic hypotension, constipation, and urinary retention. Topical medications such as capsaicin and isosorbide nitrate may be considered in the second-or third-line treatment. Opioids may not be recommended as a first-or second-line treatment acute exacerbations since it is addictive(3, 8). Numerous treatments are palliative in nature and do not target the underlying mechanism causing the pain. Specifically, alpha-lipoic acid and epalrestat appear to modify disease state well by modulating pathogenesis but are not recommend to by guidelines(8). Therefore, there is an urgent need to discover new therapeutic agents that can target the mechanisms.

Traditional Chinese medicine (TCM) has a unique theoretical system. It has accumulated rich clinical experience in the treatment of T2DM and related complications(9). The concept of DPN is not clearly described in TCM. Based on the characteristics of “numbness, pain, coldness and limb weakness”, DPN belongs to the categories of “Xiaoke (emaciation and thirst)”, “Xiaoke pulse Bi”, “Bi syndrome” and “Wei syndrome” in the concept of TCM(6, 10). Mudan granules is a Chinese patent medicine consisting of Astragalus membranaceus (Huangqi), Corydalis rhizoma with vinegar (Yanhusuo), Panax notoginseng (Sanqi), Radix Paeoniae rubra (Chishao), Salvia miltiorrhiza (Danshen), Ligusticum chuanxiong, safflower (Honghua), Logwood (Sumu), and Caulis Spatholobi (Jixueteng)(11). It has the effects of invigorating Qi and promoting blood circulation, unblocking the meridians, and relieving pain. In China it is approved for the treatment of DPN(6). A randomised controlled trial including 148 patients with DPN showed that Mudan granules improved the conduction velocity of the common peroneal and median nerves and were effective in relieving clinical symptoms(12). Another study showed that Mudan granules combined with pancreatic kininogenase increased serum SOD level, decreased serum hypersensitive C-reactive protein (hs-CRP) level and TCSS score, and improved sensory nerve function in DPN patients(13). Therefore, in this systematic review, we evaluated clinical randomized controlled trials (RCTs) and pre-clinical studies of Mudan granules combined with RT for DPN, in order to more objectively evaluate the efficacy and safety of Mudan granules and to explore the potential mechanisms.

## 2. Methods

### 2.1. Search strategy

The meta-analysis followed the guidelines for Preferred Reporting Items for Systematic Reviews and Meta-analyses (PRISMA)(14) (see **S1 File** for the PRISMA checklist). The study was registered on the PROSPERO platform (CRD42022373113). The literature search was not restricted by language. The literature search was conducted using combinations of the following words: “diabetic peripheral neuropathy or diabetic or neuralgia” and “Mudan or Mu Dan”. Electronic databases, including PubMed, Embase, Cochrane, China National Knowledge Infrastructure (CNKI), VIP database, Wanfang database were searched. The detailed search strategies were provided in the additional file (see **S2 File**). In addition, we manually searched the reference lists of included clinical trials. The retrieval time range was from the establishment of the database to October 2022. Two authors (Jianlong Zhou and Lv Zhu) independently conducted the literature search based on the search strategy. Disputed sections were resolved by the third author (Yadi Li).

### 2.2. Eligible criteria

The inclusion criteria were as follows: (I) the study participants were clinically diagnosed with DPN; (II) the control group was treated as follows: routine treatment (e.g., diet modification, regular exercise, blood glucose control, etc.) combined or not with western medicine for the treatment of DPN; (III) Patients in the trial group were treated with Mudan granules on the basis of treatment in the control group; (IV) the study design is RCTs or preclinical studies; (V) there is no restriction on the duration of disease or treatment; (VI) outcome measures included at least one of effectiveness and safety measure; effectiveness measures included total clinical efficacy (TCE), nerve conduction velocity (NCV) including at least one of the median, common peroneal and tibial nerves, TCSS score, TSS, serum homocysteine (Hcy) level, serum hs-CRP level, serum SOD level; safety measure included adverse event rate.

The exclusion criteria were as follows: (I) duplicate literature; (II) review; (III) case report; (IV) clinical experience; (V) data analysis; (VI) review; (VII) non-diabetic peripheral neuropathy; (VIII) unrelated intervention; (IX) protocol or guideline; (X) non-randomised controlled trial; (XI) lack of data.

### 2.3. Study screening and Date extraction

We used reference management software (EndNote 20) to screen the literature. First, duplicates were identified and removed. Then, the literature was screened based on title and abstract. Finally, the full text was reviewed for eligibility based on inclusion and exclusion criteria. Two researchers (JLZ and LZ) independently assessed all retrieved literature and analysed the data according to inclusion/exclusion criteria. Disagreements were resolved by the third researcher (YDL). The relevant information was extracted and recorded using Microsoft excel software. The following information was extracted: (1) basic information of the included studies, including year of publication, name of first author, country or region, number of cases per group, baseline characteristics of subjects (age); (2) intervention and course of treatment in the trial and control groups; (3) outcome measures, including: total clinical efficacy (TCE), nerve conduction velocity (NCV) including at least one of the median, common peroneal and tibial nerves, TCSS score, TSS, serum homocysteine (Hcy) level, serum hs-CRP level, serum SOD level, adverse event. Data will be cross-checked by the third researcher to ensure quality.

### 2.4. Quality assessment

The study quality was also evaluated independently by two researchers (ZJL and ZL) according to the risk of bias assessment tool recommended by the Cochrane Systematic Review Manual (5.1.0). The evaluation included the following 7 items: random sequence generation; allocation concealment; blinding of participants and personnel; blinding of outcome assessment; incomplete outcome data; selective reporting; other sources of bias. Three levels of risk of bias were followed to assess: low risk, unclear risk, and high risk. Risk of bias plots for the evaluation results were drawn using the R package.

### 2.5. Data analysis

Stata 16.0 software (Stata Corporation, College Station, Texas, USA) was used for the meta-analysis of all data. For dichotomous data, we calculated risk ratio (RR) and 95% confidence interval (CI). Standardized mean difference (SMD) and 95% CI were calculated for continuous data. The statistical significance of differences between groups was evaluated with the Z-test, and *P* <= 0.05 indicated that the differences were statistically significant. The random-effect model of meta-analysis was used in the study, assuming significant heterogeneity in all the included studies. Cochran’s Q test and Higgins’ *I*^2^ test were used to analyze the heterogeneity between studies. When *P* > 0.05 or *I*^2^ < 50%, there is no significant heterogeneity between studies. Otherwise, it means there exists significant heterogeneity among studies. In addition, Funnel plots were used to assess the publication bias.

L’Abbe plot were also used to test for heterogeneity.

### 2.6. Patient and public involvement

Patients and public were not involved in this study.

## 3. Results

### 3.1. Literature search

A total of 179 articles were retrieved according to the search strategy. After removing duplicates, 152 articles potentially relevant to this study were retained. Among these, 52 articles were excluded after initial screening of titles and abstracts. After reading the full text of the remaining 100 studies, 56 studies were included. All studies were conducted in China, with 51 clinical studies and 5 preclinical studies. Ultimately, 51 clinical studies were included in the meta-analysis(12, 13, 15–63). The flowchart of the retrieval is shown in **Fig 1**.

**Fig 1.**
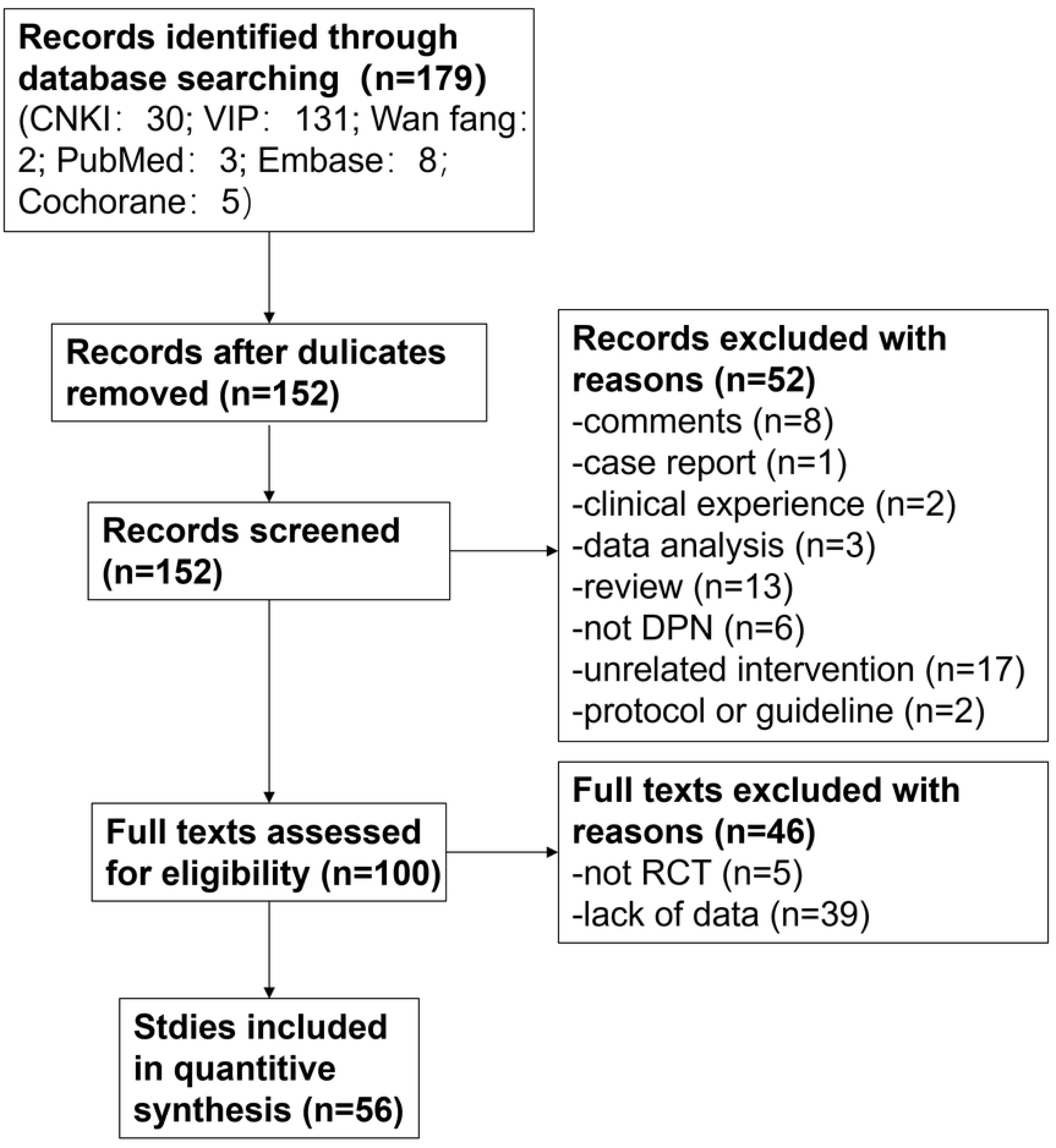
Flow chart of literature retrieval and inclusion.

### 3.2. Characteristics

A total of 51 studies were included in this study for quantitative analysis, involving 5416 patients, including 2752 in the trial group and 2664 in the control group. The control group was treated as follows: RT (e.g., diet modification, regular exercise, blood glucose control, etc.) combined or not with RT for DPN. Patients in the trial group were treated with Mudan granules based on treatment in the control group. The basic information of the included literature is shown in **S1 Table**.

### 3.3. Methodological quality

The quality of the 51 included studies was assessed using the RCT risk of bias assessment tool recommended by the Cochrane Collaboration. The results of the risk of bias assessment are shown in **Figs 2** and **3**. Three studies grouped patients according to drug regimens(37, 45, 60) and two studies grouped patients according to order of admission(49, 50), all of whom were rated as high risk. 25 studies explicitly stated that patients were grouped according to rolling dice or random number table methods, all of which were rated as low risk. Studies that did not specify the method of randomization were rated as unclear risk. All randomized controlled trials without allocation concealment and blinding were rated as unclear risk. All included studies showed a low risk of bias in terms of selective reporting and incomplete outcome data. Three studies were rated as high risk in term of other sources of bias because the mean age of patients enrolled was not reported(22, 25, 55).

**Fig 2.**
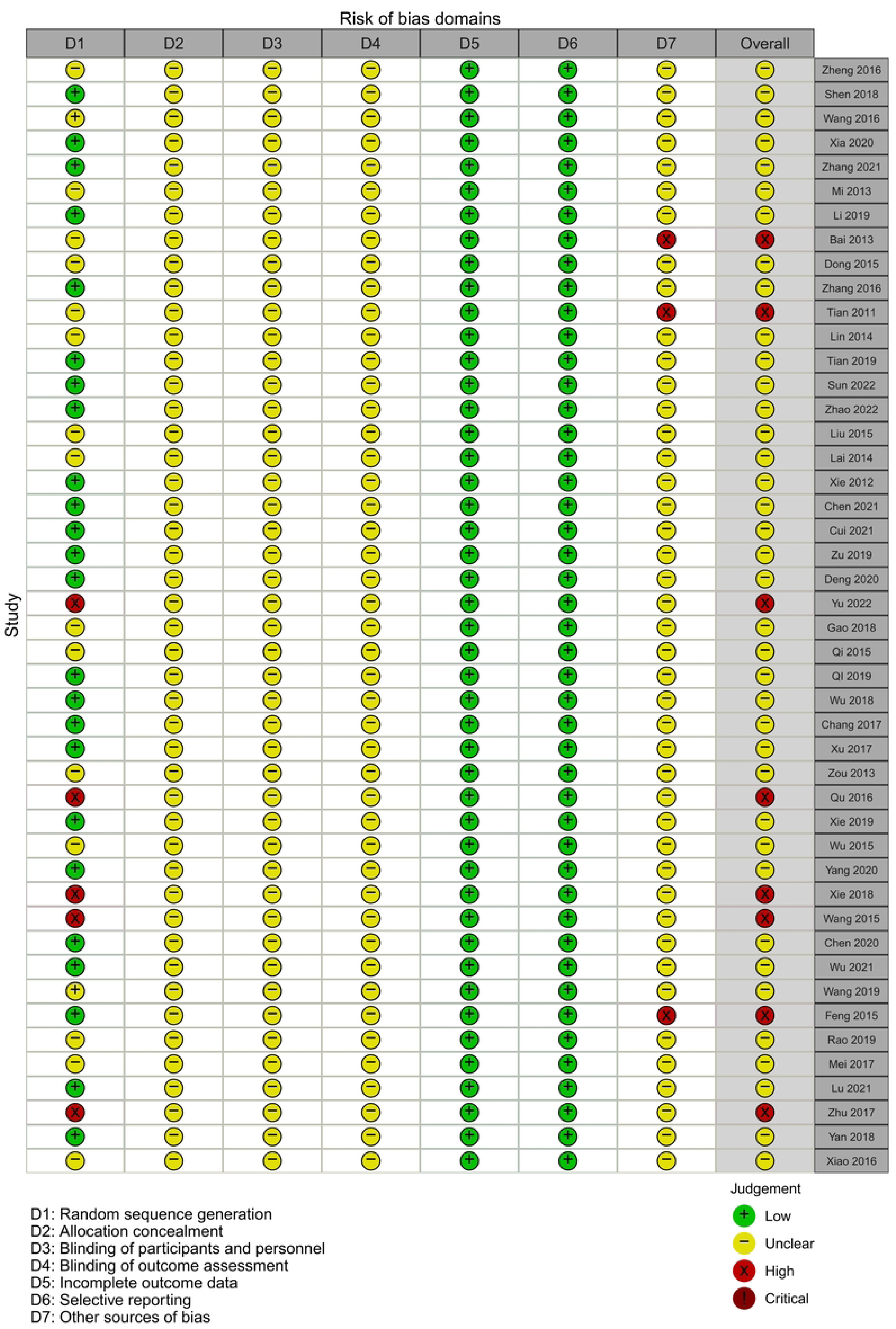
Risk of bias assessment of the included literature.

**Fig 3.**
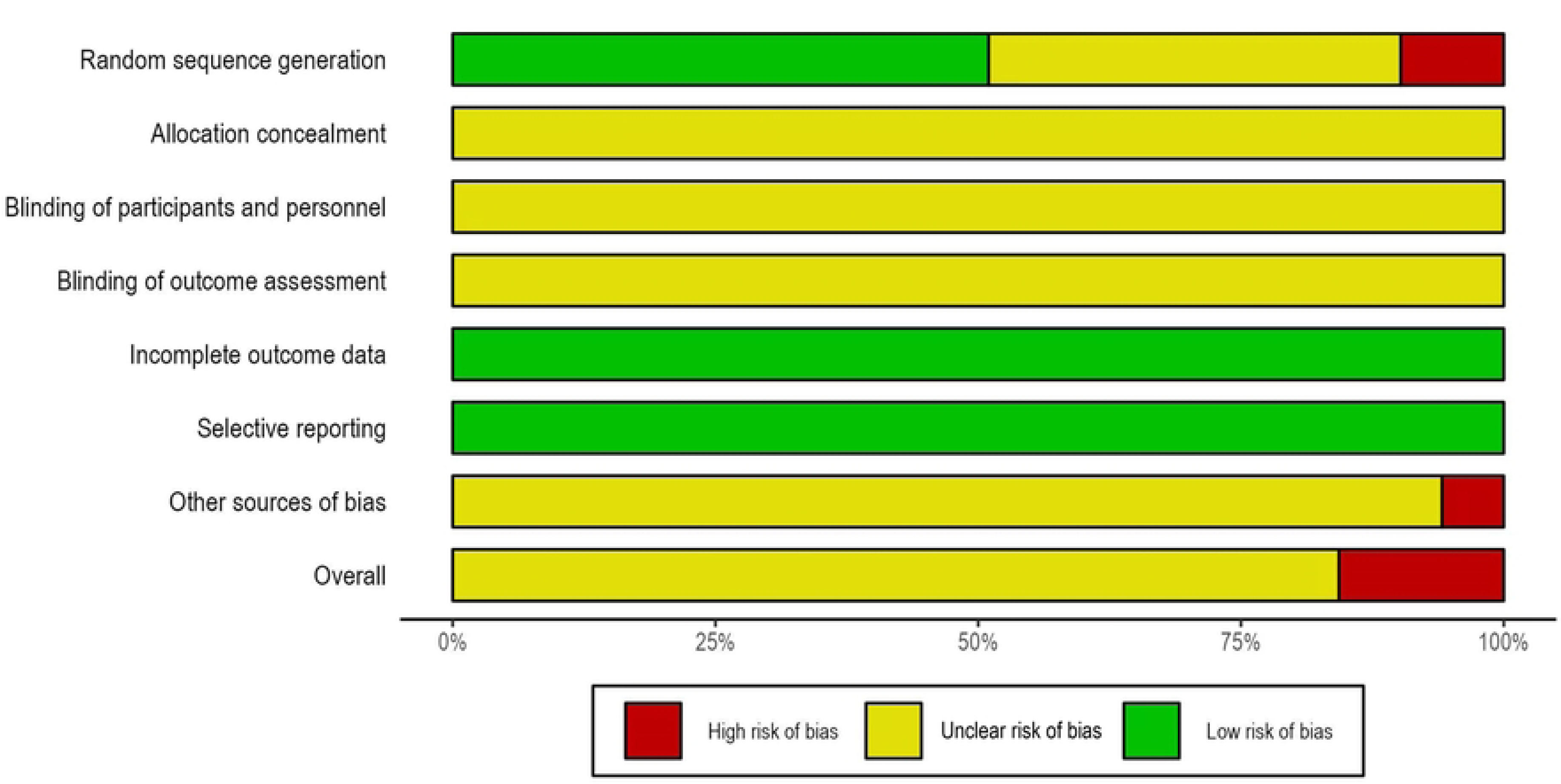
Bar chart of risk of bias assessment for the included literature.

### 3.4. Meta-analysis

#### 3.4.1. Total clinical efficacy (TCE)

A total of 41 articles reported TCE, and there was no significant heterogeneity between studies (*I*^2^ =20%, *P* =0.26). The results of the meta-analysis indicated that the TCE of combining Mudan granules plus RT were 1.23 times (RR 1.23, 95% CI: 1.19 to 1.27, *P* <0.01) more effective than the RT for DPN, as shown in **Fig 4**. We also performed a heterogeneity test for TCE using the L’Abbe plot. The L’Abbe plot showed that all studies were above the null line and that the studies showed a more concentrated distribution. This also indicated that the efficacy of Mudan granules for DPN was superior to that of the control group, as shown in **Fig 5**.

**Fig 4.**
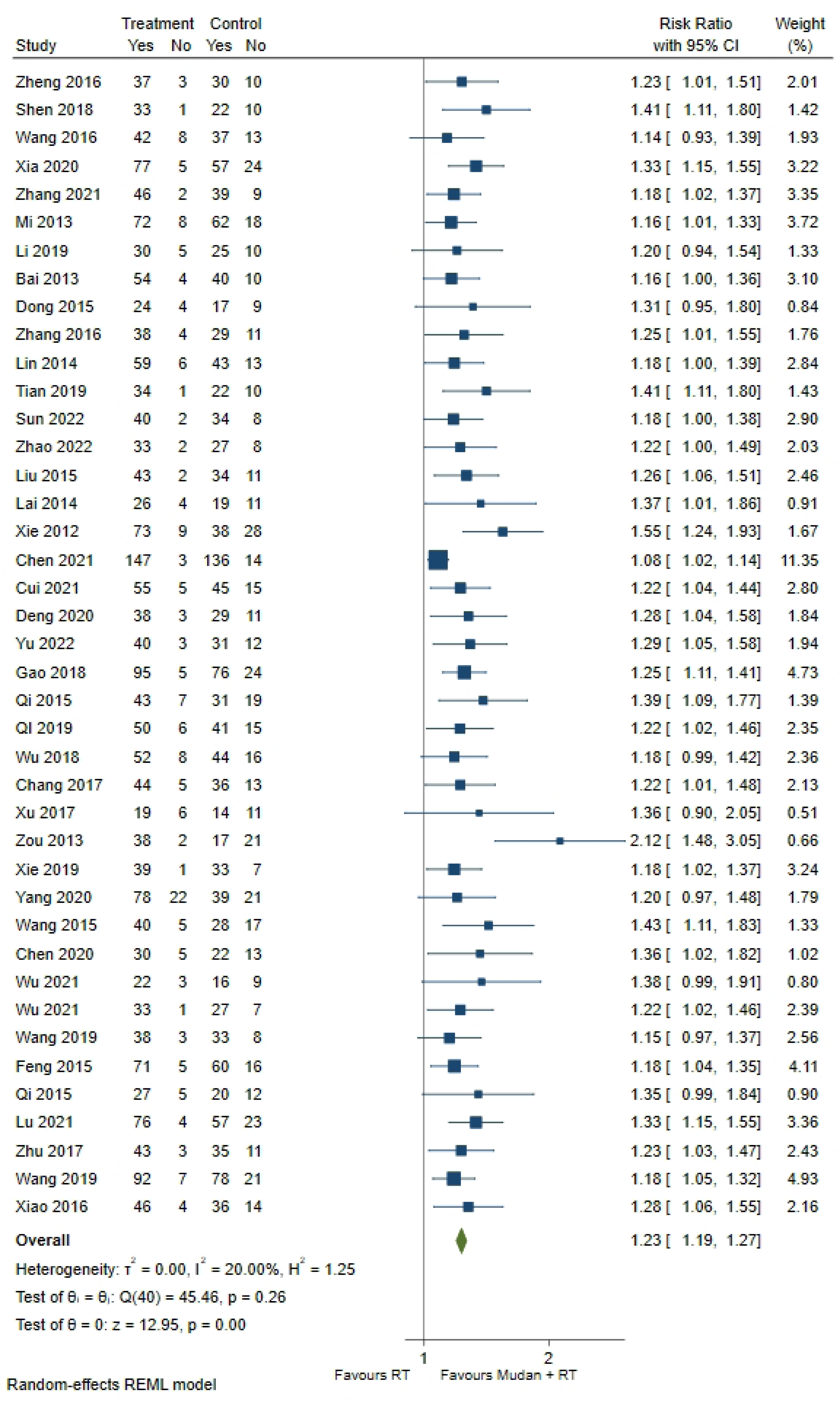
Forest plot of total clinical efficacy.

**Fig 5.**
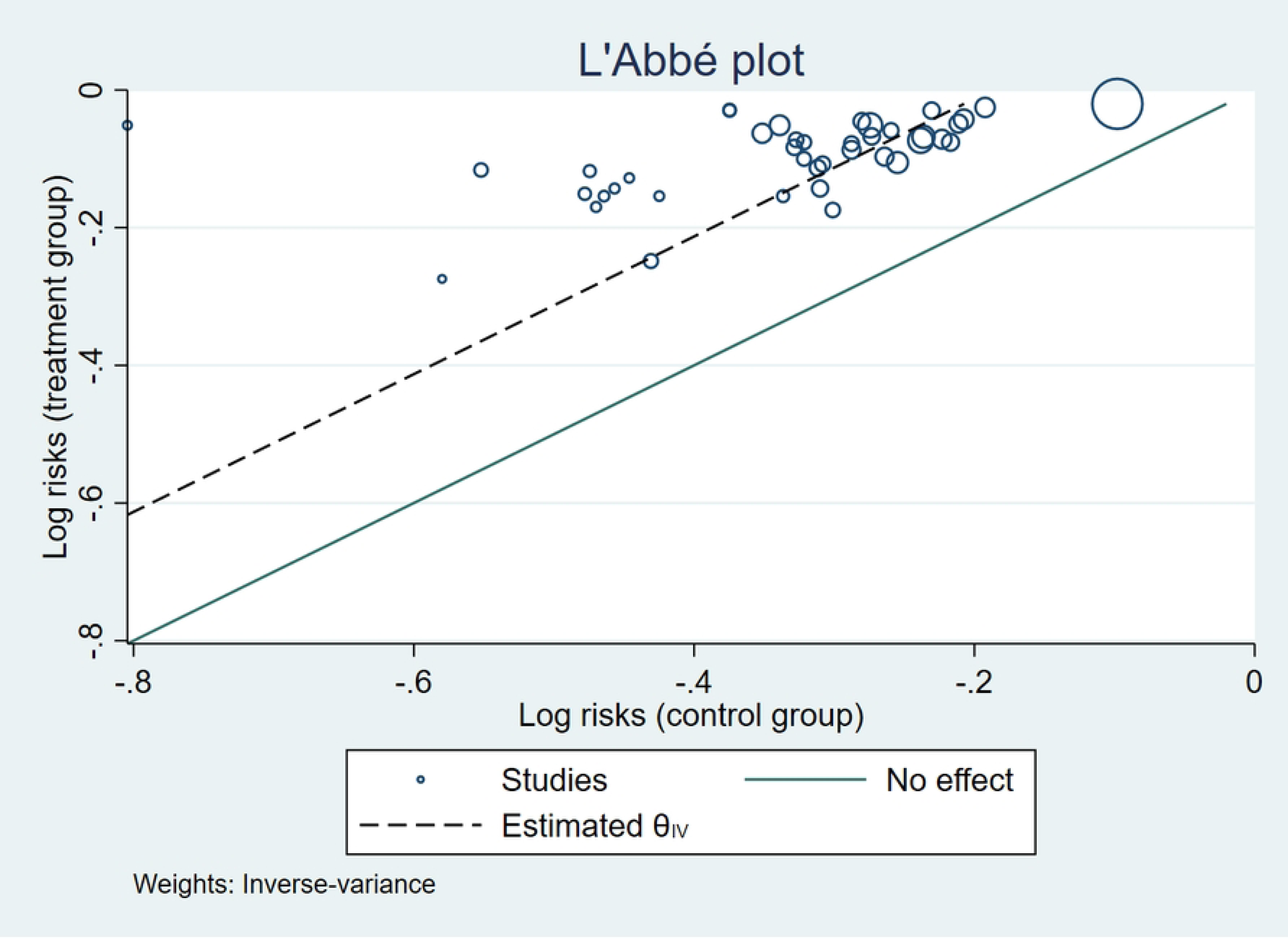
L’Abbe plot of heterogeneity test for total clinical efficacy.

#### 3.4.2. Median nerve conduction velocity

A total of 29 articles reported changes in median motor nerve conduction velocity before and after treatment. SMD (i.e., Cohen’s d) was estimated using a random effect model. As shown in **Table 1**, the difference in median motor nerve conduction velocity before and after treatment with Mudan granules based on the treatment of the control group for DPN was statistically significant (SMD 1.61, 95% CI: 1.16 to 2.07, *P* <0.01). There was a high heterogeneity between studies (*I*^2^ =96.07%, *P* <0.01). A total of 27 articles reported changes in median sensory nerve conduction velocity before and after treatment. As shown in **Table 1**, the difference in median sensory nerve conduction velocity before and after treatment with Mudan granules was statistically significant compared with the control group (SMD 1.73, 95% CI: 1.26 to 2.20, *P* <0.01), and there was significant heterogeneity between studies (*I*^2^ =96.19%, *P* <0.01). The above results suggested that Mudan granules were effective in increasing median nerve conduction velocity of patients with DPN. The detailed results were shown in **S1 Fig**.

**Table 1.** Meta-analysis results of other outcome measures.

| Outcome measures | Pooled studies | Analysis mode | $I^2$ | Effect size | Pooling value with 95% CI | $P$ |
| --- | --- | --- | --- | --- | --- | --- |
| Median nerve MNCV | 29 | Random effect model | 96.07% | SMD | 1.61[1.16, 2.07] | <0.01 |
| Median nerve SNCV | 27 | Random effect model | 96.19% | SMD | 1.73[1.26, 2.20] | <0.01 |
| Common peroneal nerve MNCV | 38 | Random effect model | 96.34% | SMD | 1.48[1.10, 1.86] | <0.01 |
| Common peroneal nerve SNCV | 35 | Random effect model | 95.15% | SMD | 1.57[1.23, 1.92] | <0.01 |
| Tibial never MNCV | 8 | Random effect model | 92.73% | SMD | 1.34[0.82, 1.87] | <0.01 |
| Tibial never SNCV | 5 | Random effect model | 0% | SMD | 1.03[0.86, 1.20] | <0.01 |
| TCSS score | 9 | Random effect model | 0% | SMD | -0.52[-0.66, -0.38] | <0.01 |
| TSS | 5 | Random effect model | 97.44% | SMD | -1.44[-2.88, -0.00] | 0.05 |
| **Serum Hcy level** | 5 | Random effect model | 98.42% | SMD | -3.84[-5.99, -1.70] | <0.01 |
| **Serum hs-CRP level** | 11 | Random effect model | 99.39% | SMD | -1.68[-3.29, -0.08] | 0.04 |
| **Serum SOD level** | 15 | Random effect model | 92.44% | SMD | 1.54[1.13, 1.95] | <0.01 |
| **Adverse event** | 20 | Random effect model | 0% | RR | 0.96[0.63, 1.48] | 0.87 |

*Note.* MNCV, Motor nerve conduction velocity; SNCV, Sensory nerve conduction velocity; TCSS, Toronto clinical scoring system; TSS, Total symptom score; hs-CRP, Hypersensitive C-reactive protein; SOD, Superoxide dismutase; Hcy, Homocysteine.

#### 3.4.3. Common peroneal nerve conduction velocity

A total of 38 articles reported changes in common peroneal motor nerve conduction velocity before and after treatment. As shown in **Table 1**, the difference in common peroneal motor nerve conduction velocity before and after treatment with Mudan granules based on the treatment of the control group for DPN was statistically significant (SMD 1.48, 95% CI: 1.10 to 1.86, *P* <0.01). There was significant heterogeneity between studies (*I*^2^ =96.34%, *P* <0.01). A total of 35 articles reported changes in common peroneal sensory nerve conduction velocity before and after treatment. Compared with the control group, there was a statistically significant difference in the conduction velocity of the common peroneal sensory nerve before and after treatment with Mudan granules (SMD 1.57, 95% CI: 1.23 to 1.92, *P* <0.01), and there was significant heterogeneity between studies (*I*^2^ =95.15%, *P* <0.01). The above results suggested that Mudan granules were effective in increasing the common peroneal nerve conduction velocity in DPN patients. The detailed results were shown in **S2 Fig**.

#### 3.4.4. Tibial nerve conduction velocity

A total of 8 studies reported changes in tibial nerve motor nerve conduction velocity before and after treatment. As shown in **Table 1**, the difference in tibial nerve motor nerve conduction velocity before and after treatment with Mudan granules was statistically significant based on the treatment of DPN in the control group (SMD 1.34, 95% CI: 0.82 to 1.87, *P* <0.01). There was significant heterogeneity between studies (*I*^2^ =92.73%, *P* <0.01). A total of 5 articles reported changes in tibial nerve sensory nerve conduction velocity before and after treatment. The difference in tibial nerve sensory nerve conduction velocity before and after treatment with Mudan granules was statistically significant compared with the control group (SMD 1.03, 95% CI: 0.86 to 1.20, *P* <0.01), and there was no heterogeneity between studies (*I*^2^ =0%, *P* =0.39). These results suggested that Mudan granules could effectively improve tibial nerve conduction velocity in DPN patients. The detailed results were shown in **S3 Fig**.

#### 3.4.5. Toronto Clinical Scoring System (TCSS) score

A total of 9 studies reported changes in the TCSS score before and after treatment. The results of the meta-analysis showed that there was a statistically significant difference in TCSS score before and after treatment with Mudan granules compared with the control group (SMD −0.52, 95% CI: −0.66 to −0.38, *P* <0.01). There was a low heterogeneity between studies (*I*^2^ =0%, *P* =0.76), as shown in **Table 1** and **S4 Fig**. This suggested that Mudan granules may effectively reduce the TCSS score of DPN patients.

#### 3.4.6. Total Symptom Score (TSS)

A total of 5 articles reported changes in TSS before and after treatment. There was a high heterogeneity between studies (*I*^2^ =97.44%, *P* <0.01). The difference in TSS before and after treatment with Mudan granules was statistically significant compared to the control group (SMD - 1.44, 95% CI: −2.88 to −0.00, *P* =0.05). This indicated that Mudan granules could effectively reduce TSS in DPN patients. As shown in **Table 1** and **S5 Fig**.

#### 3.4.7. Serum Hcy level

A total of 5 articles reported changes in serum Hcy level before and after treatment. Compared with RT, serum Hcy level was significantly lower in Mudan granules plus RT treatment (SMD −3.84, 95% CI: −5.99 to −1.70, *P* <0.01), and there was significant heterogeneity between studies (*I*^2^ =98.42%, *P* <0.01). As shown in **Table 1** and **S6 Fig**.

#### 3.4.8. Serum hs-CRP level

A total of 11 studies reported the changes in serum hs-CRP level before and after treatment. Compared with RT, serum hs-CRP level was significantly lower in Mudan granules plus RT treatment (SMD −1.68, 95% CI: −3.29 to −0.08, *P* =0.04), and there was significant heterogeneity between studies (*I*^2^ =99.39%, *P* <0.01). As shown in **Table 1** and **S7 Fig**.

#### 3.4.9. Serum SOD level

A total of 15 studies reported changes in serum SOD level before and after treatment. Compared with RT alone, serum SOD level was significantly higher in Mudan granules plus RT treatment (SMD 1.54, 95% CI: 1.13 to 1.95, *P* <0.01), and there was a high heterogeneity between studies (*I*^2^ =92.44%, *P* <0.01). As shown in **Table 1** and **S8 Fig**.

#### 3.4.10. Adverse event

A total of 20 studies reported the adverse events, with a low heterogeneity between studies (*I*^2^ =0%, *P* =0.89). The difference in the rate of adverse event after Mudan granules treatment was not statistically significant compared with the control group (RR 0.96, 95% CI: 0.63 to 1.48, *P* =0.87), as shown in **Table 1** and **S9 Fig**. Both had low rates of adverse events, and no serious adverse events were reported.

#### 3.4.11. Sensitivity analysis

For outcome measures without significant statistical heterogeneity, the results of the random effect model were compared with those of the fixed effect model, and the results showed that the analysis was stable and reliable.

#### 3.4.12. Publication bias analysis

We performed a funnel plot analysis of total clinical efficacy. The funnel plot showed an uneven distribution of studies, which implies a possible publication bias, as shown in **Fig 6**. The following problems existed in the literature analysis: the large number of studies with low sample size, poor methodological design of the studies, etc. All the above factors may have caused the funnel plot to show an uneven distribution.

**Fig 6.**
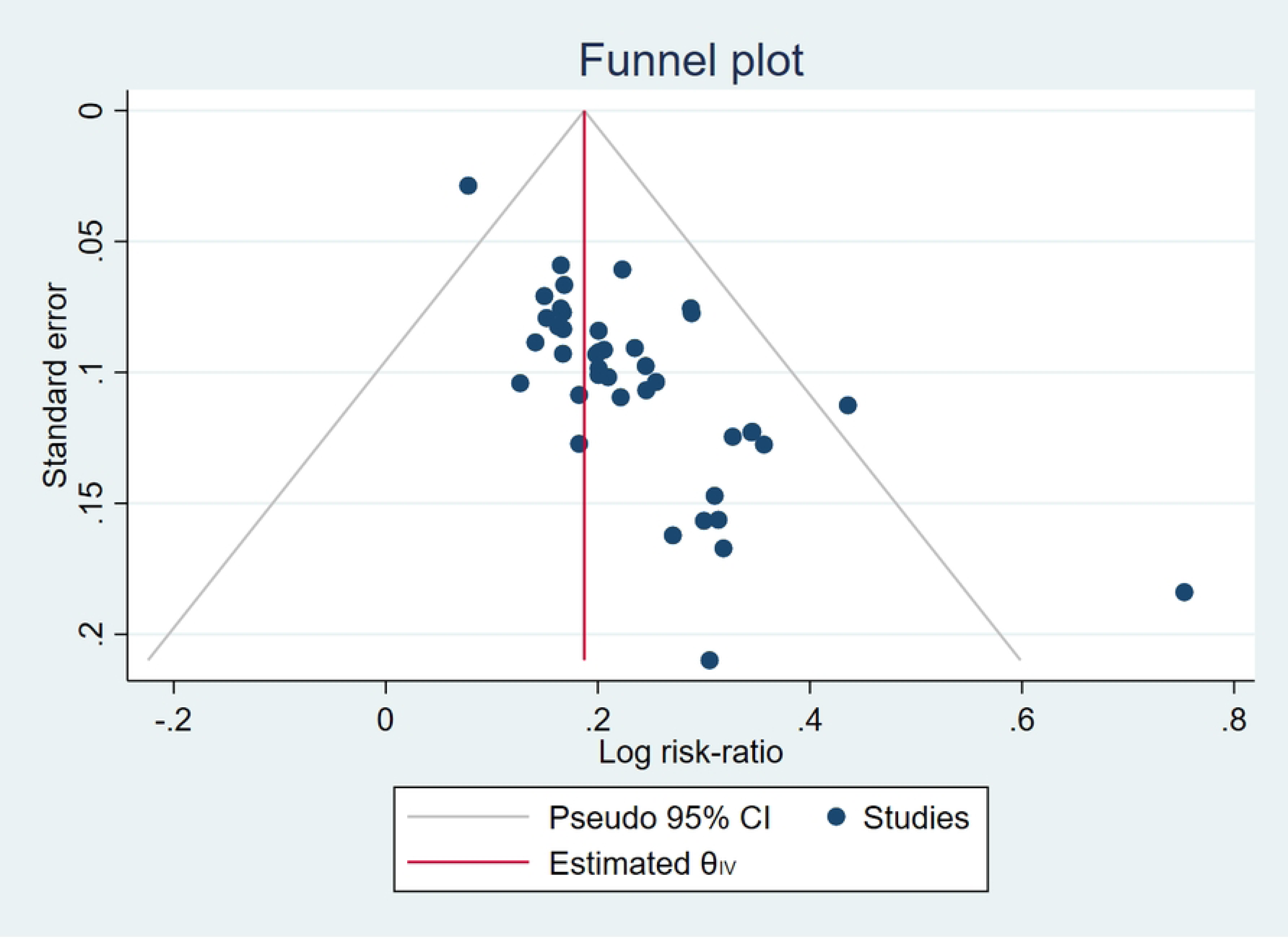
Funnel plot of the total clinical efficacy of the two groups of patients.

#### 3.4.13. Analysis of the results of preclinical studies

We included 5 preclinical studies. After analyzing these articles, we summarized the possible mechanisms of Mudan granules for DPN, as shown in **Fig 7**. Animal experiments have shown that Mudan granules could improve sciatic nerve conduction velocity(64) and reduce inflammatory response by decreasing serum Hcy and TGF-β1 levels(65). Mudan granules are also able to increase serum glutathione (GSH) and SOD levels and reduce serum malondialdehyde (MDA) levels(66), which indicated its ability to resist oxidative stress. Several studies have also found that Mudan granules could improve peripheral nerve injury by modulating the TLR4/MyD88/NF-κB pathway and the TLR4/p38 MAPK pathway(67). It may also alleviate neuropathic pain in diabetic rats by modulating the PI3K/AKT signaling pathway(68).

**Fig 7.**
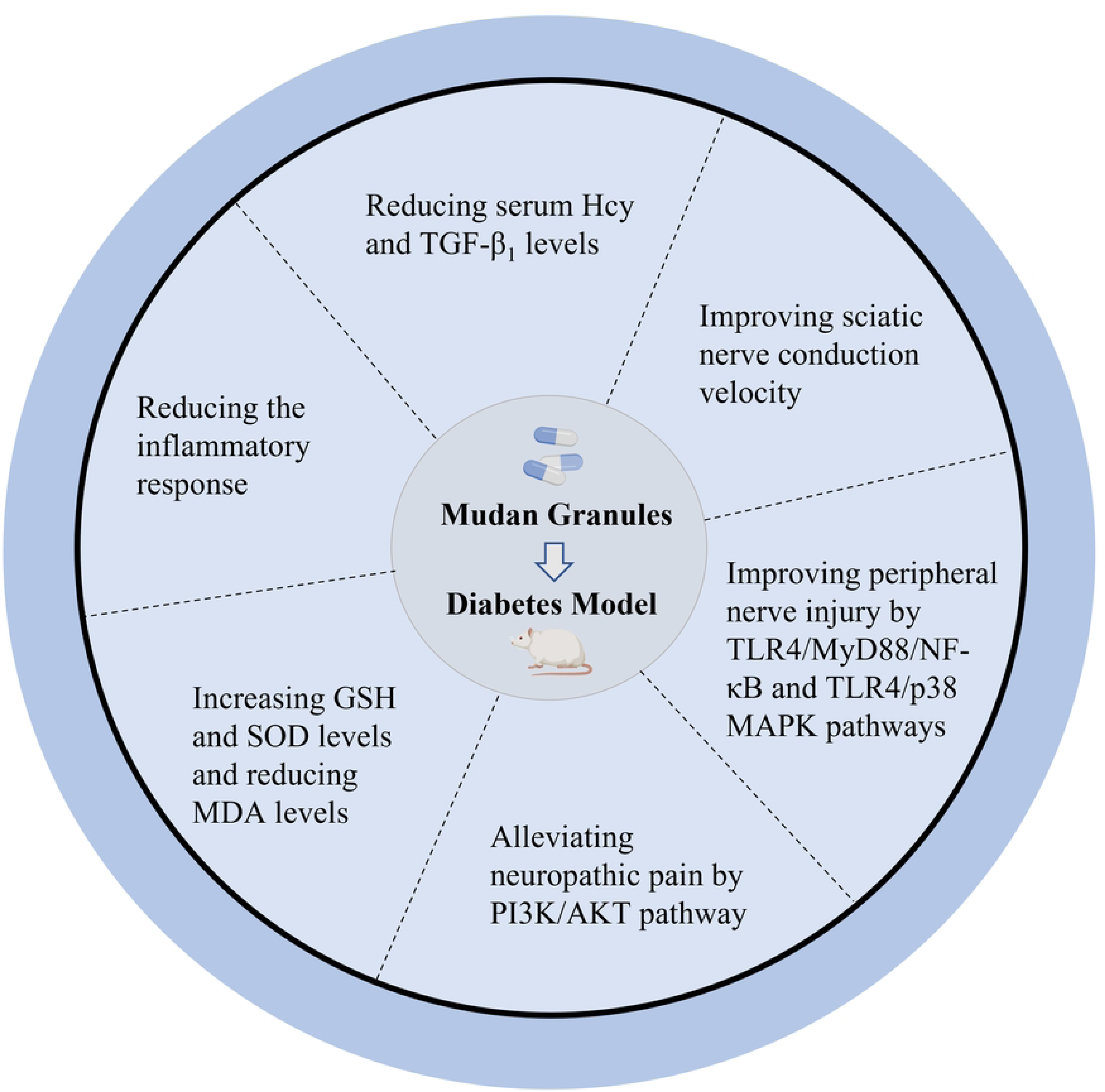
Mechanistic plot of Mudan granules in the treatment of DPN.

## 4. Discussion

In the study, we evaluated the efficacy and safety of Mudan granules in patients with DPN and explored the relevant mechanisms. Previous study included 11 RCTs and found that Mudan granules had a significant advantage over RT alone for DPN in terms of overall efficiency and improvement in the conduction velocity of the common peroneal, median, and tibial nerves. Its conclusion was that Mudan granules was effective and safe in the treatment of DPN(69). Our study also showed that Mudan granules was effective and safe in the treatment of DPN, with results consistent with previous meta-analysis. However, the main problems with these early studies were the limited sample size and the small number of outcome measures. Our study included studies from 2017 onwards and added outcome indicators such as TCSS score, TSS, Hcy, hs-CRP, and SOD. This study showed that compared with the control group, Mudan granules was able to reduce TCSS score and TSS, lower serum Hcy and hs-CRP levels, and increase SOD level.

Except for the low heterogeneity of outcome measures such as total clinical efficacy, TCSS score, tibial sensory nerve conduction velocity, and adverse event, the heterogeneity of Meta-analysis was greater for other measures, which may be related to the following two reasons. First, the Western medicine treatment measures were inconsistent between studies. For example, in some studies, the control group was treated with mecobalamin in addition to routine treatment, while in others it was combined with lipoic acid, and in still others, the control group was treated with epalrestat or other drugs. Second, the duration of therapy varied between studies. Some studies had a treatment duration of 2 weeks, while some studies had a treatment duration of 4 weeks, and others had a treatment duration of 12 weeks.

There are several limitations to this meta-analysis that need to be addressed. First, despite the inclusion of RCTs, the main studies included had large biases in the methodological design, for example, all studies did not specify allocation concealment and blinding. Only 25 studies provided sufficient information about the randomization process. Second, for most of the studies, the treatment duration was not long enough to properly assess the efficacy of Mudan granule treatment. This is because DPN is a chronic disease that requires long-term treatment. The long-term efficacy and safety studies are essential to determine the therapeutic utility of the drug. However, the duration of treatment in the included studies ranged from 1 to 12 weeks, and long-term follow-up studies may yield different results. Therefore, we were unable to assess the long-term safety and efficacy of Mudan granules for the treatment of DPN. Third, study sample sizes were generally small, and most studies did not involve formal prior sample size calculations. Hence, the use of large samples in RCTs is needed to further validate the findings and avoid overestimation of intervention benefits due to insufficient sample size. Finally, although we retrieved studies published in English or Chinese databases, the language of the final included studies was Chinese, and studies published from other languages were missing. Our findings may not be well disseminated.

The pathogenesis of DPN is still not fully elucidated. It is commonly believed that DPN is associated with hyperglycemia, hyperlipidemia, insulin resistance and protein metabolism(5). Some studies have found that serum Hcy level was closely related to the development of DPN(70). High Hcy levels can influence the function of insulin sensitivity, oxidative stress, nitric oxide, and the pathways of vascular and neurological damage(71). Studies have shown that serum CRP levels were found to be an independent predictor of the presence of DPN(72). Elevated serum TNF-α and IL-10 and CRP is a phenomenon associated with a pro-inflammatory response to diabetes, which was also associated with diabetic neuropathy(73). Oxidative stress caused by hyperglycemia was considered to be the initiating factor in the pathogenesis of DPN(74). Some other studies have shown that SOD is an important antioxidant enzyme. In DPN rats, serum SOD was decreased, while serum MDA was significantly increased(74). Our study found that Mudan granules were able to reduce serum Hcy, hs-CRP and MDA levels and increase serum SOD levels, which showed its anti-inflammatory and anti-oxidative stress abilities. Some studies have found that insulin deficiency or insulin resistance caused insulin receptors to promote apoptosis via the P13K/AKT pathway(6). Defective endoneurial microvasculature may lead to hypoxia and ischemia, generation of oxidative stress, and activation of the redox-sensitive transcription factor NFκB(5). Our study found that Mudan granules may improve peripheral nerve injury by modulating the TLR4/MyD88/NF-κB pathway and TLR4/p38 MAPK pathway and alleviate neuropathic pain in diabetic rats by modulating the PI3K/AKT signaling pathway. Other studies have found that the potential mechanisms of Mudan granules for DPN may include improving insulin resistance, reducing islet cell apoptosis, neuroprotection, and other effects(11, 75). Increasing evidence has suggested(76–78) that hyperlipidemia contributes to the progression of diabetic neuropathy. Total cholesterol, triglycerides, and low-density lipoprotein (LDL) cholesterol have been proved to be associated with DPN. A study on Mudan granules for diabetic dorsalis pedis atherosclerosis showed that Mudan granules could reduce serum total cholesterol, triglycerides, and LDL cholesterol levels(79). This predicted that Mudan granules may treat DPN by regulating lipid metabolism, and further studies on the lipid metabolism-related pathways of Mudan granules for DPN could follow. These studies above showed that Mudan granules may improve the development of DNP through a multi-target and multi-pathway approach.

## 5. Conclusions

Our systematic and meta-analytic study provides supportive evidence that Mudan granules are effective in the treatment of DPN patients based on RT, and no serious adverse events were observed. High-quality studies are essential to be conducted and further prove the preliminary evidence from the preclinical studies and clinical studies. Additionally, prolonged observation and follow-up are also needed to demonstrate the long-term efficacy and safety of Mudan granules.

## Data Availability

All relevant data are within the manuscript and its Supporting Information files.

## Data Availability

All relevant data are within the manuscript and its Supporting Information files.

## Acknowledgements

None.

## Authors’ Contributions

Jianlong Zhou: Conceived and designed the study, drawn up the manuscript and analysed the data, prepared figures and/or tables. Lv Zhu: collected the data and performed the analysis. Yadi Li: supervised and revised the manuscript. All authors read and approved the manuscript.

## Supporting information

**S1 File. PRISMA checklist.**

**S2 File. The search strategies of English databases.**

**S1 Table. The basic information of the included literature.**

**S1 Fig. (A) Forest plot of the improvement of median motor nerve conduction velocity by Mudan granules. (B) Forest plot of the improvement of median sensory nerve conduction velocity by Mudan granules.**

**S2 Fig. (A) Forest plot of Mudan granules improving the common peroneal motor nerve conduction velocity. (B) Forest plot of Mudan granules improving the common peroneal sensory nerve conduction velocity.**

**S3 Fig. (A) Forest plot of Mudan granules improving the tibial motor nerve conduction velocity. (B) Forest plot of Mudan granules improving the tibial sensory nerve conduction velocity.**

**S4 Fig. Forest plot of Mudan granules for reducing TCSS score.**

**S5 Fig. Forest plot of TSS reduction by Mudan granules.**

**S6 Fig. Forest plot of Mudan granules reducing serum Hcy level.**

**S7 Fig. Forest plot of Mudan granule reducing serum hs-CRP level.**

**S8 Fig. Forest plot of elevated serum SOD level by Mudan granules.**

**S9 Fig. Forest plot comparing the rate of adverse events in the two groups.**

## Reference

1. Carmichael J, Fadavi H, Ishibashi F, Shore AC, Tavakoli M. Advances in Screening, Early Diagnosis and Accurate Staging of Diabetic Neuropathy. Front Endocrinol. 2021;12:25.

2. Selvarajah D, Kar D, Khunti K, Davies MJ, Scott AR, Walker J, et al. Diabetic peripheral neuropathy: advances in diagnosis and strategies for screening and early intervention. Lancet Diabetes Endocrinol. 2019;7(12):938–48.

3. Hicks CW, Selvin E. Epidemiology of Peripheral Neuropathy and Lower Extremity Disease in Diabetes. Curr Diabetes Rep. 2019;19(10):8.

4. Pop-Busui R, Boulton AJM, Feldman EL, Bril V, Freeman R, Malik RA, et al. Diabetic Neuropathy: A Position Statement by the American Diabetes Association. Diabetes Care. 2017;40(1):136–54.

5. Burgess J, Frank B, Marshall A, Khalil RS, Ponirakis G, Petropoulos IN, et al. Early Detection of Diabetic Peripheral Neuropathy: A Focus on Small Nerve Fibres. Diagnostics. 2021;11(2):39.

6. Yang K, Wang Y, Li YW, Chen YG, Xing N, Lin HB, et al. Progress in the treatment of diabetic peripheral neuropathy. Biomed Pharmacother. 2022;148:10.

7. Khdour MR. Treatment of diabetic peripheral neuropathy: a review. J Pharm Pharmacol. 2020;72(7):863–72.

8. Rafiullah M, Siddiqui K. Pharmacological Treatment of Diabetic Peripheral Neuropathy: An Update. CNS Neurol Disord-Drug Targets. 2022;21(10):884–900.

9. Zheng WJ, Wang GF, Zhang Z, Wang ZG, Ma K. Research progress on classical traditional Chinese medicine formula Liuwei Dihuang pills in the treatment of type 2 diabetes. Biomed Pharmacother. 2020;121:7.

10. Piao YL, Liang XC. Chinese Medicine in Diabetic Peripheral Neuropathy: Experimental Research on Nerve Repair and Regeneration. Evid-based Complement Altern Med. 2012;2012:13.

11. Zhang YH, Jin D, Duan YY, Hao R, Chen KY, Yu TY, et al. Efficacy of Mudan Granule (Combined With Methylcobalamin) on Type 2 Diabetic Peripheral Neuropathy: Study Protocol for a Double-Blind, Randomized, Placebo-Controlled, Parallel-Arm, Multi-Center Trial. Front Pharmacol. 2021;12:10.

12. Xie X. Randomized parallel controlled study of treating type 2 diabetic peripheral neuropathy by Mudan particles plus westem medicine. J Pract Tradit Chin Intern Med. 2012;26(05):53–4 (in Chinese).

13. Sun J. Mudan granule and kallidinogenase in treatment of DPN patients and the influence on SOD and hs-CRP. Liaoning J Tradit Chin Med. 2022;49(03):127–30 (in Chinese).

14. Page MJ, McKenzie JE, Bossuyt PM, Boutron I, Hoffmann TC, Mulrow CD, et al. The PRISMA 2020 statement: an updated guideline for reporting systematic reviews. BMJ. 2021;372:n71.

15. Zheng M, Du YB, Chen BQ, Wei GZ, Zhao Y. Clinical study of antioxidant stress of integrated traditional Chinese and western medicine on diabetic peripheral neuropathy. Yunnan J Tradit Chin Med Materia Med. 2016;37(02):30–1 (in Chinese).

16. Shen YD, Wu JF. Clinical study on kallidinogenase, Mudan granule and meeobalamin in treatment of diabetic neuropathy. Diabetes New World. 2018;21(22):14–7 (in Chinese).

17. Wang YN, Yu SJ. Clinical observation of Mudan graule on the treatment of diabetic peripheral neuropathy. J Liaoning Univ Tradit Chin Med. 2016;18(6):87–9 (in Chinese).

18. Xia XH, Yan LH, Wang HD. Effectiveness of Mudan granules in the treatment of patients with diabetic peripheral neuropathy. Henan Med Res. 2020;29(24):4557–9 (in Chinese).

19. Zhang JJ, Cao W, Zhao YJ, Hu B, Wu YP. The curative effect of Mudan granule in the treatment of diabetic peripheral neu-ropathy and its influence on serum oxidative stress,inflammatory factors and pain substance levels. World J Integr Tradit West Med. 2021;16(12):2265–70 (in Chinese).

20. Mi YX, Mi YX, Zhou FJ. Clinical research on Mudan Granule in treating diabetic peripheral neuropathy. Journal of Hunan Univ of CM. 2013;33(11):65–7.

21. Li J, Fan FW, Tu Y. Clinical efficacy and safety of mudan granules in the treatment of diabetic peripheral neuropathy. Chin J Clin Rati Drug Use. 2019;12(09):70–1 (in Chinese).

22. Bai HC. Clinical observation of Mu Dan particles to treat diabetic peripheral neuropathy. World Notes on Antibiotics. 2013;34(03):133–4 (in Chinese).

23. Dong M, Wen ZJ, Song YQ, Chen J. The efficacy of Mudan granules in the treatment of diabetic peripheral neuropathy. Liaoning J Tradit Chin Med. 2015;42(07):1278–9 (in Chinese).

24. Zhang J, Qiu SL, Liang YH, Jin M. Clinical efficacy of Mudan granules in the treatment of diabetic peripheral neuropathy with Qi deficiency and blood stasis. Beijing J Tradit Chin Med. 2016;35(12):1166–8 (in Chinese).

25. Tian H, Y., Zhang ZG. Comparison of the efficacy of mudan granules and micropowder in the treatment of diabetic peripheral neuropathy. Mod J Integr Tradit Chin West Med. 2011;20(25):3166–7 (in Chinese).

26. Lin JH. Clinical observation of Mu Dan granule combined with mecobalamin in the treatment of diabetic peripheral neuropathy. Chinese Community Doctors. 2014;30(20):101–2 (in Chinese).

27. Tian W. Efficacy of the Mudan granule plus conventional western medicine on diabetic peripheral neuropathy and its electromyography. Clin J Chin Med. 2019;11(05):74–6 (in Chinese).

28. Zhao D. Impact of Mudan granule plus insulin on nerve conduction velocity of diabetic peripheral neuropathy patients. Liaoning J Tradit Chin Med. 2022;49(01):116–9 (in Chinese).

29. Wang LN, Du J, Wen ZJ, Yan Y. Efficacy of mudan granules combined with epalrestat in the treatment of diabetic peripheral neuropathy. Liaoning J Tradit Chin Med. 2016;43(09):1890–1 (in Chinese).

30. Liu L, Qin GJ. Efficacy of mudan granules combined with epalrestat in the treatment of diabetic peripheral neuropathy in the elderly and the effect on homocysteine. Chin J Pract Med. 2015;42(01):85–6 (in Chinese).

31. Lai J, Wang LF, Zeng JR. A comparative study on the clinical efficacy of Mudan granules combined with western medicine in the treatment of diabetic peripheral neuropathy. Asia-Pacific Traditional Medicine. 2014;10(03):99–100 (in Chinese).

32. Chen D. Effect of Mudan granules combined with calcium hydroxybenzene sulfonate in the treatment of patients with diabetic peripheral neuropathy. Medical Journal of Chinese People’s Health. 2021;33(10):85–7 (in Chinese).

33. Cui M, Peng YP, Gao JQ, Li S, Yang XS. Effects of Mudan granule combined with calcium dobesilate capsules on nerve conduction velocity,hemorheology and oxidative stress in patients with diabetic peripheral neuropathy. Pro Mod Biomed. 2021;21(22):4270–4 (in Chinese).

34. Zu Q, Xu BX. Effect of Mudan granules combined with alprostadil on nerve conduction velocity in patients with diabetic peripheral neuropathy. Chin J Rati Drug Use. 2019;16(10):12–4 (in Chinese).

35. Zhang YP, Peng XF, He F, Mao YL, Cai SY. Effect of Mudan particles combined with methylcobalamin and lipoic acid for diabetic peripheral neuropa-thy. Journal of North China University of Science and Technology Health Sciences Edition. 2016;18(06):454–7 (in Chinese).

36. Deng XJ, Ren HS. Effect of Mudan granule and thiocyanic acid injection on inflammation response and hemodynamic index in patients with diabetic peripheral neuropathy. The Medical Forum. 2020;24(16):2234–7 (in Chinese).

37. Yu L, Zhang W, Fu WT. Effects of Mudan granule combined with lipoic acid injection on TSS score and serum levels of SOD and Hcy in diabetic peripheral neuropathy patients. Sichuan J Physiol Sci. 2022;44(01):30–3 (in Chinese).

38. Gao HP. Mudan granules combined with lipoic acid and methylcobalamin for diabetic peripheral neuropathy. Pract Clin J Integr Tradit Chin West Med. 2018;18(02):19–21 (in Chinese).

39. Qi Y, Yu SJ. The clinical effect of Mudan particles combined with mecobalamine in the treatment of painful diabetic peripheral neuropathy. World Chinese Medicine. 2015;10(03):356–8 (in Chinese).

40. Qi JZ, Li L, Wang XM, Bai XP. Clinical efficacy of mudan granules combined with methylcobalamin in the treatment of diabetic peripheral neuropathy. J Med Theor & Prac. 2019;32(11):1678–9 (in Chinese).

41. Wu WY. Effect of Mudan granule combined with mecobalamin for the treatment of diabetic peripheral neuropathy and the impact on nerve conduction velocity. Clin Med Eng. 2018;25(08):1071–2 (in Chinese).

42. Chang C, Li Y. Clinical study of Mudan granule combined with Mecobalamin in treatment of diabetic peripheral neuropathy. Chin J Gen Pract. 2017;15(05):792–5 (in Chinese).

43. Xu T, Hao LM, Zhang Y, Liu CC, Liu HX. The effect of Mudan granule combined with macobalamine in the treatment of diabetic peripheral neuropathy. World Chinese Medicine. 2017;12(02):266–8 (in Chinese).

44. Zou H, Zhang G. Clinical observation on the treatment of type 2 diabetic peripheral neuropathy with Mudan granules combined with methylcobalamin. J Xinxiang Med Coll. 2013;30(09):759–60.

45. Qu Y, Zhang YP. Study of Mudan grabule combined with mecobalamin and a-lipoic acid in the treatment of diabetic peripheral neuropathy. Clin Res Pract. 2016;1(20):22–3 (in Chinese).

46. Xie YQ. Effect of Mudan granules, mecobalamine and α-lipoic acid in oxidative stress for diabetic peripheral neuropathy patients. Liaoning J Tradit Chin Med. 2019;46(10):2109–11 (in Chinese).

47. Wu XF, Luo XH, Xu J, Xu RY, Hou HB. Effect of MuDan particles combined with cobalt amine and α-lipoic acid on diabetic peripheral neuropathy. Chin J Gen Pract. 2015;13(12):1966–7+2029(in Chinese).

48. Yang LM, Jia RM, Gao Q, Yi JD, Li YZ. Therapeutic effect of Mudan granule combined with Mecobalamin on DPN (syndrome of deficiency of Qi)and collaterals and its influ-ence on neuroelectrophysiology and oxidative stress index. J Clin Exp Med. 2020;19(02):181–4 (in Chinese).

49. Zhou MQ. Efficacy of Mudan particles with methylcobalamin and gabapentin on patients with painful diabetic peripheral neuropathy. Liaoning J Tradit Chin Med. 2016;43(11):2317–9 (in Chinese).

50. Wang X, Cui HM, Ren F, Kong C, He Y. Effectiveness of mudan granules combined with methylcobalamin and alpha-lipoic acid in the treatment of diabetic peripheral neuropathy. Clin Med Chin. 2015;31(02):124–6 (in Chinese).

51. Chen Y. Effects of Mudan granule combined with insulin glargine on painful diabetic peripheral neuropathy. Chin J Pract Med. 2020;47(15):108–11 (in Chinese).

52. Wu Y. Effect of Mudan granule combined with routine western medicine on nerve conduction velocity and its curative effect on diabetic peripheral neuropathy. Liaoning J Tradit Chin Med. 2021;48(01):112–5 (in Chinese).

53. Wu HJ, Shi F, Zhang Y, Song XJ, Wang YL. Study on the effect of Mudan granule combined with beraprost sodium and epalrestat in the treatment of diabetic peripheral neuropathy. Diabetes New World. 2021;24(05):177–8+85 (in Chinese).

54. Wang LN, Du J, Dong M, Wen ZJ. Effect of Mudan granules, beraprost sodium tablets and epalrestat on diabetic peripheral neuropathy. Liaoning J Tradit Chin Med. 2019;46(11):2354–7 (in Chinese).

55. Feng CE, Shi ZD. Efficacy of Mudan particles combined beraprost and epalrestat for diabetic peripheral neuropathy. Evaluation and Analysis of Drug-Use in Hospitals of China. 2015;15(01):44–6 (in Chinese).

56. Rao XJ, Wu YM. Clinical therapeutic observation of Mudan granule plusα-lipoic acid in patients with diabetic peripheral neuropa-thy. Chin J Pract Nerv Dis. 2019;22(04):370–3 (in Chinese).

57. Mei LN, Wu CA. 46 cases of diabetic peripheral neuropathy treated with mudan granules combined with alpha-lipoic acid. Zhejiang J Tradit Chin Med. 2017;52(01):70 (in Chinese).

58. Qi Y, Yu SJ. Effect of mudan granules on oxidative stress in painful diabetic peripheral neuropathy. Lishizhen Med Mater Med Res. 2015;26(7):1561–3 (in Chinese).

59. Lu L, Wu L. Evaluation of the efficacy of Mudan granules in diabetic peripheral neuropathy. Chin Remed Clin. 2021;21(13):2294–5 (in Chinese).

60. Zhu ZQ, Cheng SJ. Effectiveness of methylcobalamin combined with Mudan granules in the treatment of diabetic peripheral neuropathy. Mod Pract Med. 2017;29(04):503–4 (in Chinese).

61. Wang Y, Liu L. Effectiveness of methylcobalamin combined with mudan granules in the treatment of diabetic peripheral neuropathy. Women’s Health Research. 2019(19):123–4 (in Chinese).

62. Wen BH, Shen SM. Comparison of the curative effect of different regimens on diabetic peripheral neuropathy. J Hunan Norm Univ (Med Sci). 2018;15(03):81–4 (in Chinese).

63. Xia CJ. Effect of combined treatment of diabetic peripheral neuropathy with α-lipoic acid and Mudan granules on Hcy and clinical effects. Chin Foreign Med Res. 2016;14(09):61–3 (in Chinese).

64. Yang WQ, Yu YB, Xu XL, Ren HX, Liang JF, Zhang L. Efficacy of Mu Dan granules on experimental rats with diabetic peripheral neuropathy. Chin J Diabetes. 2015;23(12):1099–102 (in Chinese).

65. Sun YM, Zhao HH, Yin YL, Wang L. Effect of Mudan granule on Hcy and TGF-β1 in serum of rat with diabetic peripheral neuropathy in different course of disease. Shanxi J of TCM. 2021;37(10):51–3 (in Chinese).

66. Li YY, Yu SJ. The Effect of Mudan Particles on the Oxidative Levels of Spinal Cord of Diabetic Rats. Medical Recapitulate. 2016;22(10):2070–3 (in Chinese).

67. Zhou M, Yao WJ. Study on Regulatory Effects of Mudan Granules on TLR4-related Pathway in DPN Rats. Western J of TCM. 2021;34(10):52–6 (in Chinese).

68. Qi Y, Liu YT, Zhang H, Zhao ZJ. Research of Mudan granule improved neuropathic pain in diabetic rats by regulating PI3K/AKT pathway. Chin J Diffic and Compl Cas. 2021;20(02):176–81 (in Chinese).

69. Fang ZH, Chen XQ, Chen YM, Qiu JJ, Yang C. Meta-analysis of the efficacy of Mudan granules in the treatment of diabetic peripheral neuropathy. Journal of New Chinese Medicine. 2017;49(08):154–8 (in Chinese).

70. Lin XY, Sun T, Yu RX, Tang YL, Li HY. Correlation between elevated serum homocysteine level and the development of diabetic peripheral neuropathy: a comparative study and meta-analysis. Int J Clin Exp Med. 2016;9(2):2857–64.

71. Zhao JD, Li Y, Xin L, Sun M, Yu CJ, Shi GB, et al. Clinical Features and Rules of Chinese Herbal Medicine in Diabetic Peripheral Neuropathy Patients. Evid-based Complement Altern Med. 2020;2020:8.

72. Canpolat AG, Emral R, Keskin C, Canlar S, Sahin M, Corapcioglu D. Association of monocyte-to-high density lipoprotein-cholesterol ratio with peripheral neuropathy in patients with Type II diabetes mellitus. Biomark Med. 2019;13(11):907–15.

73. Nadi M, Bambaeichi E, Marandi SM. Comparison of the effect of two therapeutic exercises on the inflammatory and physiological conditions and complications of diabetic neuropathy in female patients. Diabetes Metab Syndr Obes. 2019;12:1493–501.

74. Zhang YZ, Zhou ZC, Song CY, Chen X. The Protective Effect and Mechanism of Dexmedetomidine on Diabetic Peripheral Neuropathy in Rats. Front Pharmacol. 2020;11:11.

75. Liu S-N, Sun S-J, Liu Q, Hou S-C, Shen Z-F. Effect of Mudan Granule on islets beta cell function in monosodium glutamate induced obese mice with insulin resistance: an experimental study. Zhongguo Zhong xi yi jie he za zhi Zhongguo Zhongxiyi Jiehe Zazhi= Chinese Journal of Integrated Traditional and Western Medicine. 2014;34(7):853–8.

76. Wiggin TD, Sullivan KA, Pop-Busui R, Amato A, Sima AAF, Feldman EL. Elevated Triglycerides Correlate With Progression of Diabetic Neuropathy. Diabetes. 2009;58(7):1634–40.

77. O’Brien PD, Guo K, Eid SA, Rumora AE, Hinder LM, Hayes JM, et al. Integrated lipidomic and transcriptomic analyses identify altered nerve triglycerides in mouse models of prediabetes and type 2 diabetes. Dis Model Mech. 2020;13(2):12.

78. Naqvi S, Imani S, Hosseinifard H, Wen QL, Shahzad MN, Ijaz I, et al. Associations of serum low-density lipoprotein and systolic blood pressure levels with type 2 diabetic patients with and without peripheral neuropathy: systemic review, meta-analysis and meta-regression analysis of observational studies. BMC Endocr Disord. 2019;19(1):16.

79. Guo Y, Sun Q. Effect of Mudan granule combined with zinc sulfate on ankle brachial index in diabetic foot dorsal artery sclerosis and its efficacy. Chinese Journal of Biochemical Pharmaceutics. 2016:151–4.

